# Avidity sequencing of whole genomes from retinal degeneration pedigrees identifies causal variants

**DOI:** 10.1101/2022.12.27.22283803

**Authors:** Pooja Biswas, Adda Villanueva, Benjamin J. Krajacich, Juan Moreno, Junhua Zhao, Anne Marie Berry, Danielle Lazaro, Bryan R. Lajoie, Semyon Kruglyak, Radha Ayyagari

## Abstract

Whole genome sequencing has been an effective tool in the discovery of variants that cause rare disease. In this study, we determined the suitability of a novel avidity sequencing approach for rare disease applications. We built a sample to results workflow, combining the novel sequencing technology with standard library preparation kits, analysis workflows, and interpretation tools. We applied the workflow to ten pedigrees with inherited retinal degeneration (IRD) phenotype. Candidate variants of interest identified through whole genome sequencing were further evaluated using segregation analysis. Mutations in known IRD genes were detected in five of the ten cases. Genes with identified high confidence variants associated with retinal degeneration included *PEX6, ABCA4, CERKL, MAK*, and *RDH12*. Pending confirmatory clinical sequencing, we observed a 50% diagnostic yield, consistent with previously reported outcomes of IRD patient analysis. The study confirms that avidity sequencing is effective in detection of causal mutations when used for whole genome sequencing in rare disease applications.

## Introduction

Rare genetic diseases affect millions of people around the world (Ferreira 2019). Due to the difficulties of obtaining a definitive diagnosis in many rare disease cases, the use of whole genome sequencing (WGS) has become a popular approach to identify variants that may explain the underlying cause (Carss et al. 2017; Sanford et al. 2019; Souche et al. 2022; Stranneheim et al. 2021; Turro et al. 2020; Willig et al. 2015). WGS enables a comprehensive analysis across regions (coding and noncoding) and variant classes from SNPs to CNVs (The 100,000 Genomes Project Pilot Investigators et al. 2021). An important category of rare diseases is inherited retinal degeneration, a group of rare eye diseases affecting approximately 1 in 2,000 individuals worldwide (Sohocki et al. 2001). Retinal degeneration leads to progressive loss of vision as a result of cell death within the retina. The phenotypes, including age of onset, vary widely suggesting that the underlying genetics may also be variable. Previous publications have demonstrated that exome and genome sequencing are an effective approach for establishing a molecular diagnosis in retinal degeneration cases (Carss et al. 2017). Whole genome sequencing has been applied in a wide diversity of studies at both the cohort and population scale but nearly all of these studies have used the sequencing by synthesis technology from Illumina (Bentley et al. 2008). Assessing the performance, efficiency and relative capabilities of multiple technologies has only recently become possible as more sequencing technologies have entered the commercial market. This study applied a novel technology, termed sequencing by avidity (Arslan et al. 2022), for the genetic analysis of retinal degeneration by whole genome sequencing. Briefly, the sequencing technology works as follows: DNA of interest is fragmented, circularized, and hybridized to a flowcell. Rolling circle amplification is used to generate concatemers from each circular molecule. Following a primer hybridization, avidity sequencing proceeds by sequentially identifying nucleotides in the DNA of interest by iterating on the following steps: (1) the binding of a dye labeled polymer to identify the nucleotide and (2) the incorporation of an unlabeled but 3’ blocked nucleotide to advance along the DNA template. The current implementation of avidity sequencing is capable of generating 2×150 base pair reads and producing in excess of 1 billion read pairs per flowcell, making it a good throughput match for the WGS application. To evaluate the technology on this application, components upstream and downstream of sequencing are required. Specifically, library preparation methods that are compatible with the sequencer and analysis tools that utilize the sequencer output to call and interpret variants are needed. Here we demonstrate that avidity sequencing can be integrated with standard upstream and downstream components to create an effective workflow for the WGS rare disease application.

## Results

We developed a modular architecture shown in Figure 1 to evaluate compatibility of components within a sample to results workflow. Extracted DNA was used as input to multiple library preparation kits and final libraries from two kits (one PCR-free and one PCR+) were used in this study. Once the library was prepared and sequenced, a FASTQ file was generated and used as input to standard alignment and variant calling tools. The output of the avidity sequencing platform commercialized by Element Biosciences is a standard FASTQ file compatible with all aligner and variant caller combinations tested. We selected BWA-MEM paired with either Sentieon DNAScope (Freed et al. 2022) or Google DeepTrio (Kolesnikov et al. 2021), following a benchmarking study. The VCF file produced by variant callers was used by interpretation tools to prioritize findings. Here we primarily relied on the Franklin by Genoox interpretation engine (franklin.genoox.com), though a subset of cases were also reviewed in other interpretation tools. The workflow summarized in Figure 1 was applied to samples from ten pedigrees described in Figure 2. One of the cases was a positive control that had previously been diagnosed via WGS on another technology (Biswas et al. 2021), though the interpretation process was blinded to prior results. In the other nine cases, WGS had not been previously applied and a molecular diagnosis had not been made. For one of the pedigrees (Figure 2D), we sequenced the affected individual and the parents. For all other pedigrees, we sequenced a single affected individual. Following sequencing and analysis, candidate variants that were classified as either pathogenic or likely pathogenic were further evaluated via segregation analysis to determine whether variants segregated with the phenotype across the broader pedigree. A prioritized variant that also passed segregation analysis was deemed to be causal and referred for orthologous confirmatory sequencing in a CLIA laboratory to validate the observation and to provide results that could be shared with and considered by the clinicians managing the patients. Table 1 details the findings for cases in which causal variants were identified. For the positive control, the previously published compound heterozygote in the *PEX6* gene was identified as the top candidate. The PEX6 compound heterozygous variants consisted of a frameshift indel and a missense variant. No segregation analysis was performed on these variants because this case was a previously published positive control. In four of the cases, the variants that were prioritized for validation, segregated as expected with the phenotype. In the remaining cases, no variants were classified as either pathogenic or likely pathogenic. The variants detected in *ABCA4* (homozygous frameshift), *MAK* (homozygous missense), *RDH12* (homozygous missense), and *CERKL* (homozygous stop gain) had existing ClinVar submissions classified as pathogenic or likely pathogenic. We reviewed each variant that passed segregation analysis via the Integrative Genomics Viewer (IGV) (Robinson et al. 2011). The display is provided in Figure 3. All of the calls were supported by multiple reads and occurred in generally well-behaved genomic regions. In the case of the trio, the parents were observed to be the heterozygous carriers that passed the pathogenic allele to the homozygous proband.

**Table 1:** Variants identified for each pedigree

| Figure Index | Pedigree | Gene | Variant position | Zygosity | gnomAD Frequency | ClinVar ID | ClinGen ID | Reference |
| --- | --- | --- | --- | --- | --- | --- | --- | --- |
| A | RF.FO.30 | *PEX6* | chr6-42934533 TC>T | Het | 0.000021 | 495796 | CA3811174 | (Biswas et al. 2021) |
|  |  | *PEX6* | chr6-42932993 C>A | Het | 0.000010 | 1019386 | CA3810987 | (Biswas et al. 2021) |
| B | RF.FR.0916 | *ABCA4* | chr1-94577045 C>CTTTG | Hom | 0.0000039 | 854791 | CA26842462 | (Maggi et al. 2021) |
| C | RF.BR.0416 | *MAK* | chr6-10809049 G>A | Hom | 0.000021 | 867222 | CA3633712 | (Lew et al. 2018; Stone et al. 2017) |
| D | RF.VI164.0516 | *RDH12* | chr14-68192801 C>T | Hom | 0.000011 | 2061 | CA115315 | (Zou et al. 2019; Benayoun et al. 2009) |
| E | RF.OS.0916 | *CERKL* | chr2-182423344 G>A | Hom | 0.00033 | 2364 | CA252246 | (Tuson et al. 2004; Aleman et al. 2009) |

**Figure 1:**
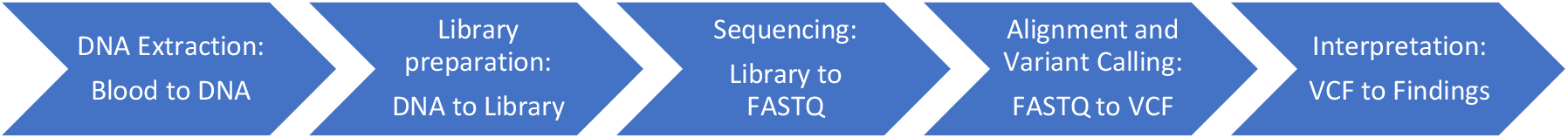
WGS for rare disease. The modular design supports multiple options for each step. In the current study, two library prep kits (with and without PCR) were used. Following avidity sequencing, Sentieon alignment and variant calling was used for single samples and DeepTrio was used for the trio. Franklin was used for interpretation with a subset of cases also reviewed via Opal.

**Figure 2:**
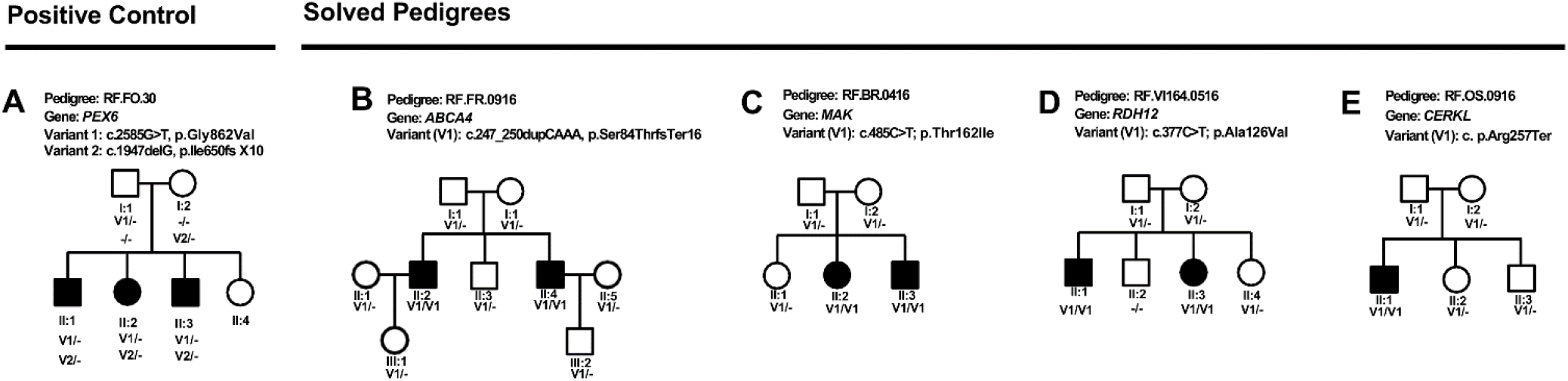
*(A) The positive control pedigree that was previously published (Biswas et al. 2021). (B)* RF.FR.0916: Two affected individuals with three unaffected individuals were recruited for the study. The whole genome analysis of individual II:2 identified a previously reported homozygous c.247_250dupCAAA p.Ser84ThrfsTer16 variant in ABCA4 gene associated with retinal degeneration (PMID-33369172). *This variant was also reported to be pathogenic by ClinVar (# 854791)*. The homozygous c.247_250dupCAAA p.Ser84ThrfsTer16 variant in ABCA4 segregated with the disease in the RF.FR.0916 pedigree. (C) *RF.BR.0416: A homozygous previously reported causal variant c.485C>T, p.Thr162Ile in MAK was identified in individual II:2 of this pedigree (ClinVar#-867222) (PMID: 28559085). The segregation analysis confirmed that this variant is present in the homozygous state in two affected individuals (II:2 and II:3) and also heterozygous in three different unaffected individuals (I:1, I:2 and II:1) including the parents (I:1 and I:2). (D) RF.VI164.0516: A family with two affected and four unaffected individuals participated in this study. A homozygous variant c.377C>T, p.Ala126Val in RDH12 gene was identified in the proband (II:1) after the whole genome analysis. This rare variant (ClinVar # 2061) segregated with the disease. (E) RF.OS.0916: Not only the proband analysis but the trio analysis of the parents and the proband from this pedigree identified a homozygous nonsense mutation p.Arg257Ter in CERKL gene (ClinVar # 2364) in the proband. We note that pedigrees are referenced via anonymized ID that cannot be used to identify patients or family members by anyone outside of the research group.*

**Figure 3:**
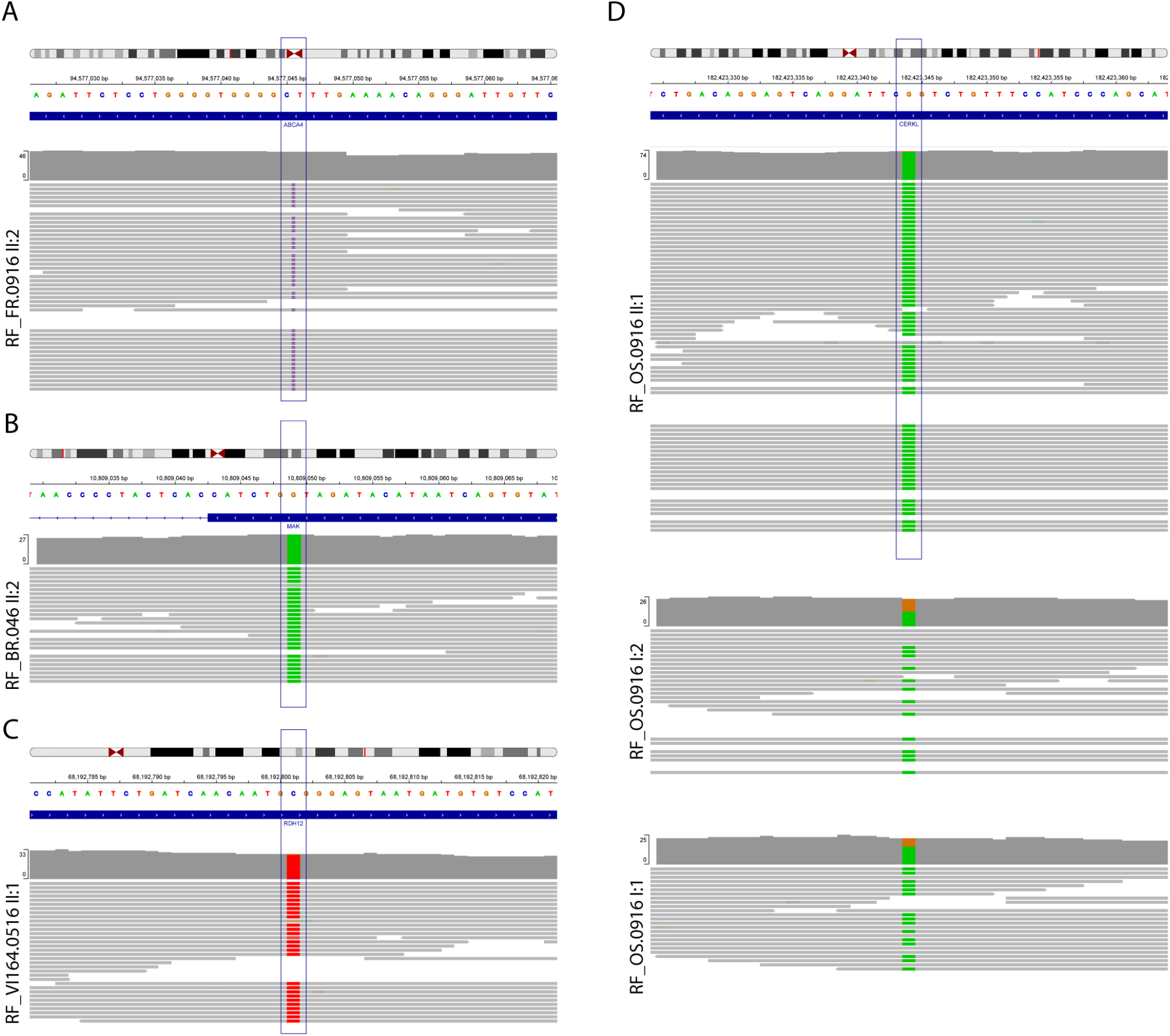
Display of IGV read alignment of the sequence encompassing the causal variants that passed segregation analysis in A. *ABCA4* (Pedigree RF.FR.0916). Coverage track shows no color because the causal variant is an insertion. B. *MAK* (RF.BR.0416), C. *RDH12* (Pedigree RF.VI164.0516). D. *CERKL* (Pedigree RF.OS.0916): read alignments in heterozygous parents I:1 and I:2 and the homozygous proband II:1. Uncolored reads in the parents support the reference allele in a heterozygous state. Deviations from 50% allele fraction may be the result of sampling variation exaggerated at reduced coverage.

## Discussion

We evaluated the new avidity sequencing technology in the context of whole genome sequencing for rare disease. To enable this evaluation, we first implemented a sample to results workflow that included library preparation, sequencing, variant calling, and variant interpretation. Our results demonstrate the success of one version of the workflow, but several alternative library preparation and analysis options exist and can be used based on laboratory preference. All samples were sequenced using 2×150 base pair reads to a target of 30X genomic coverage. For the trio, the parents were sequenced at a lower coverage so that all three samples could be combined on a single flowcell. High confidence variants were identified in five of the ten pedigrees that we sequenced, including the positive control. The variants segregated with disease through the respective pedigrees, providing support for our findings. In three pedigrees with multiple affected individuals, the sequencing of a single affected sample followed by segregation analysis resolved the entire pedigree, making this an efficient and cost-effective approach. This study demonstrates that avidity sequencing is compatible with common library preparation and analysis methods and provides an effective option for rare disease analysis using whole genome sequencing. We are in the process of expanding the patient cohort to additional retinal degeneration pedigrees and also initiating studies across other disease classes.

## Methods

### DNA Extraction from blood

Blood samples were collected from all available family members after obtaining their written consent to participate in our study. DNA extraction was done from peripheral whole blood sample, collected from all the participating patients using DNeasy Blood & Tissue Kits (Cat. No.: 69504) Qiagen (Hilden, Germany) following the manufacture’s protocol.

### Library preparation

For the singleton samples: 0.5 pmol (30 ul of 16.7nM) of pooled libraries generated with Roche KAPA HyperPlus Prep Kit (Cat# 07962428001) was processed in a single reaction using Adept Compatibility Workflow Kit (Element Biosciences, Cat# 830-00003). The final circularized library was quantified using qPCR standard and primer mix provided in the Adept Compatibility Workflow Kit following manufacturer’s protocol.

For the trio samples: DNA library preparation was performed using the combination of Roche KAPA HyperPlus Prep Kit (Roche, Cat# 07962428001) and Element Elevate Index and Adapter Kit (Element Biosciences, Cat# 830-00005), following Roche’s protocol instruction. 100ng extracted DNA was treated with enzymatic fragmentation step for 10 min, followed by end repair and A-tailing. Element Elevate adapters were used in adapter ligation step. Ligated products were purified by SPRI beads at 0.5X/0.66X ratio for size selection. 5 cycles PCR amplification was used to introduce Element index by Elevate unique index pairs (1A-1G). PCR product was cleaned up by 1x SPRI, and quantified by Qubit.

0.5 pmol (30 ul of 16.7nM) of linear library prepared above was processed in a single reaction using Element Elevate Library Circularization Kit (Cat# 830-00001). The final circularized library was quantified using qPCR standard and primer mix provided in the Elevate Library Circularization Kit following manufacturer’s protocol.

### Sequencing

The quantified libraries were pooled, denatured, and sequenced on the AVITI system using 2×150 paired end reads (Element Biosciences, San Diego CA). Two genomes were typically multiplexed per flowcell. For the trio, the three libraries were pooled on a single flowcell at unequal concentration, yielding genome coverage in the proband coverage approximately twice that of the parental samples – 52X, 20X, and 22X, respectively for proband, maternal and paternal samples.

### Analysis

Following sequencing, FASTQ files were generated as the input to secondary analysis tools. For singleton samples, the Sentieon workflow for BWA-MEM alignment and DNAScope variant calling was used. For the trio, BWA-MEM was used for alignment and Google DeepTrio was used for variant calling. The resulting variant calls were interpreted using the Franklin by Genoox software. Genoox software was also used to generate CNV candidates, though none were found in genes associated with the phenotype of interest. The phenotype was described as retinal degeneration for all cases. Franklin provided a list of 0 to 10 variants of interest for each case. In order to be further considered, variants had to be rare (<.01% gnomAD allele frequency), associated with the phenotype, and classified as likely pathogenic or pathogenic by the automated ACMG calculator. Four of the cases were also reviewed using the Opal by Fabric software (Coonrod et al. 2013) to determine if a new tool would yield additional promising candidates. If one or more variants of interest was identified, a group review was used to determine whether to proceed with segregation analysis. For the trio, the inheritance patterns were reviewed for variants of interest. For the selected variant (homozygous stop gain in CERKL), we verified that each parent was a heterozygous carrier.

### Segregation analysis

Segregation analysis of the identified variants of interest in additional family members were performed using standard PCR amplification and Sanger sequencing analysis.

### Data Access

BAM and VCF files for sequenced samples associated with the 5 pedigrees that passed segregation analysis are in the process of being submitted to dbGaP (https://www.ncbi.nlm.nih.gov/gap/).

## Data Availability

All data produced in the present study are available upon reasonable request to the authors given the restrictions of dbGAP, where the data will reside

